# A Novel Multi-ventilation Technique to Split Ventilators

**DOI:** 10.1101/2020.04.28.20083741

**Authors:** Albert Lee, Soban Umar, Nir N. Hoftman

## Abstract

**Background:** Despite efforts to initially contain Severe Acute Respiratory Syndrome Coronavirus (SARS-CoV-2), it has spread worldwide and has strained international healthcare systems to the point where advanced respiratory resources and ventilators are depleted. This study aims to explore splitting ventilators, or “multi-ventilation,” as a viable alternative in these demanding times. We investigated whether individualized tidal volume and positive end expiratory pressure (PEEP) delivery is possible to lungs of different compliances that are being simultaneously ventilated from one anesthesia ventilator.

**Methods:** We performed a controlled experiment in an operating room environment without animal or human participants. Two “test lungs” were connected to distinct modified Y-pieces that were ventilated in parallel from a single anesthesia ventilator.

**Results:** Ventilation can be manipulated to qualitatively deliver individually tailored tidal volumes in the setting of varying PEEP and compliance requirements in pressure control mode.

**Conclusions:** Splitting ventilators, or “multi-ventilation,” is a viable alternative to acute ventilator shortage during a pandemic. Ventilators can be split for individualized tidal volume and positive end-expiratory pressure delivery in multiple subjects of differing compliances and demographics.

## INTRODUCTION

In December 2019, a novel coronavirus now known as Severe Acute Respiratory Syndrome Coronavirus (SARS-CoV-2) was isolated from a cluster of patients affected in Wuhan, China. In months, despite various containment strategies, the virus had spread worldwide with almost 900,000 documented cases and over 45,000 deaths in approximately 200 countries and territories as of April 2, 2020.^1^ In the United States, despite a Society of Critical Care Medicine estimate of nearly 200,000 full-featured and basic-function ventilators, many hospital systems are already depleted of their ventilator supply.^2–4^ Though traditionally non-medical companies have offered to manufacture functional ventilators, scaling production and delivery is unlikely to be rapid enough to meet surge capacity requirements predicted in as little as one week by Institute for Health Metrics and Evaluation (IHME) estimations, drawing significant inquiry and criticism for splitting one ventilator amongst multiple patients, or “multi-ventilation.”^4,5^ Despite the joint statement from the American Society of Anesthesiologists, Society of Critical Care Medicine and American Association for Respiratory Care recommending against splitting ventilators,^6^ Surgeon General Jerome Adams and Assistant Secretary for Health Administration Brett Giroir stated multi-ventilation may be performed, but only as a last resort.^7,8^ Multiple studies have been conducted on simulation lungs or animals, but none attempted to simultaneously individualize tidal volume and positive end expiratory pressure (PEEP) in lungs of varying compliances – mainstays of treatment therapy in severe acute respiratory distress syndrome (ARDS).^9–13^ Our findings suggest that multiple patients of different compliances can be successfully ventilated from a single source with individually tailored tidal volumes and PEEP. To our knowledge, this is the first journal publication to demonstrate this.

## MATERIALS AND METHODS

Our schematic builds on past models, including the addition of flow regulation, one-way valves, and constructing a makeshift in-line PEEP valve (Figure 1). A flow sensor was unavailable at the time of simulations. All utilized parts are listed (Figure 2) and were obtained using available hospital supplies with the exception of the flow regulator, which was purchased at a hardware store. Parts were assembled as shown, with the pressure transducer attached to the elbow connector of the patient Y-piece and zeroed appropriately at the level of the Y-piece.

**Figure 1:**
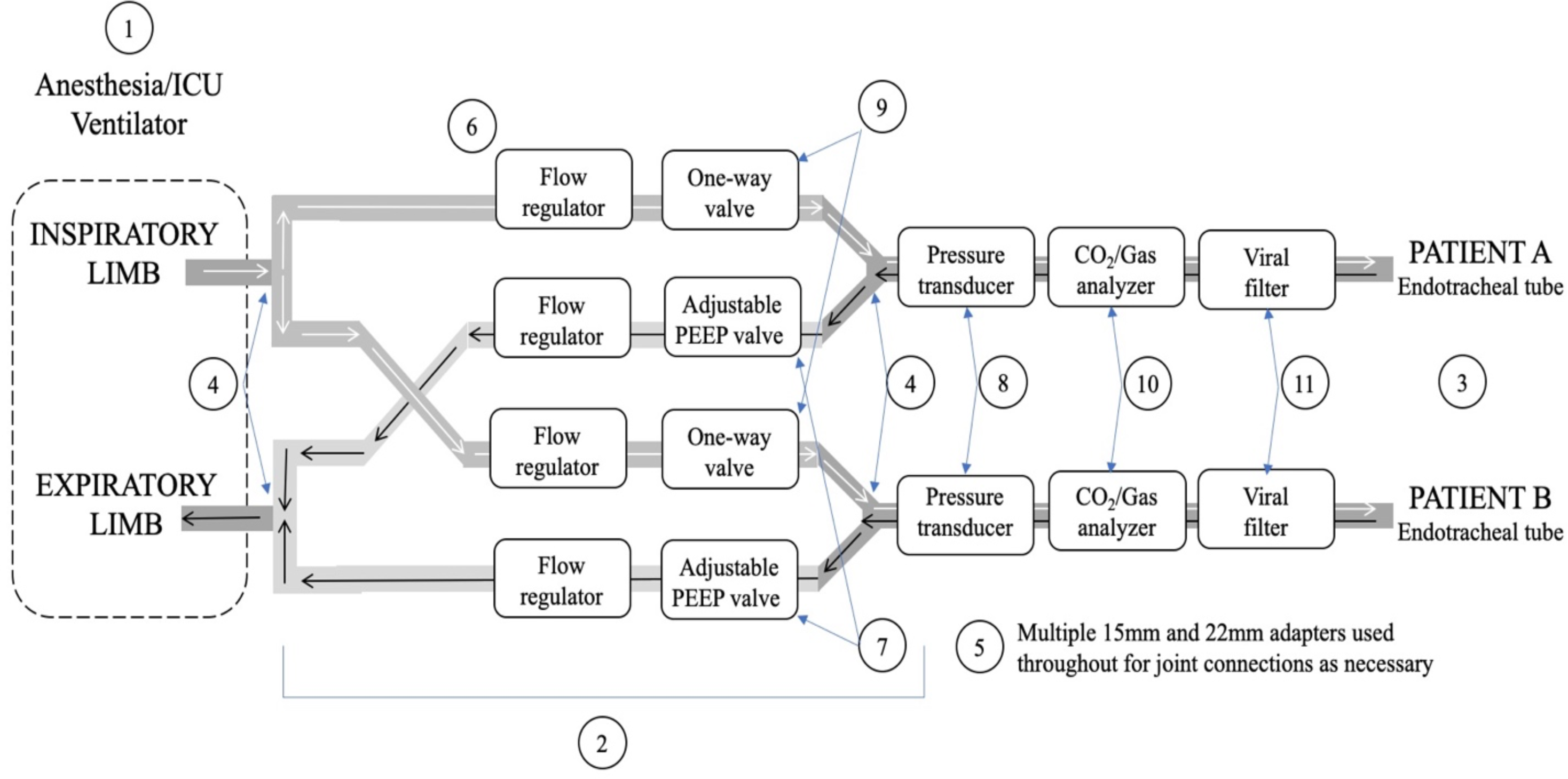
A schematic for a proposed multi-ventilation model is shown. White and black arrows indicate direction of airflow to and from the patient (white: inspiratory flow; black: expiratory flow). Encircled numbers correspond to parts used that are listed in Figure 2.

**Figure 2:**
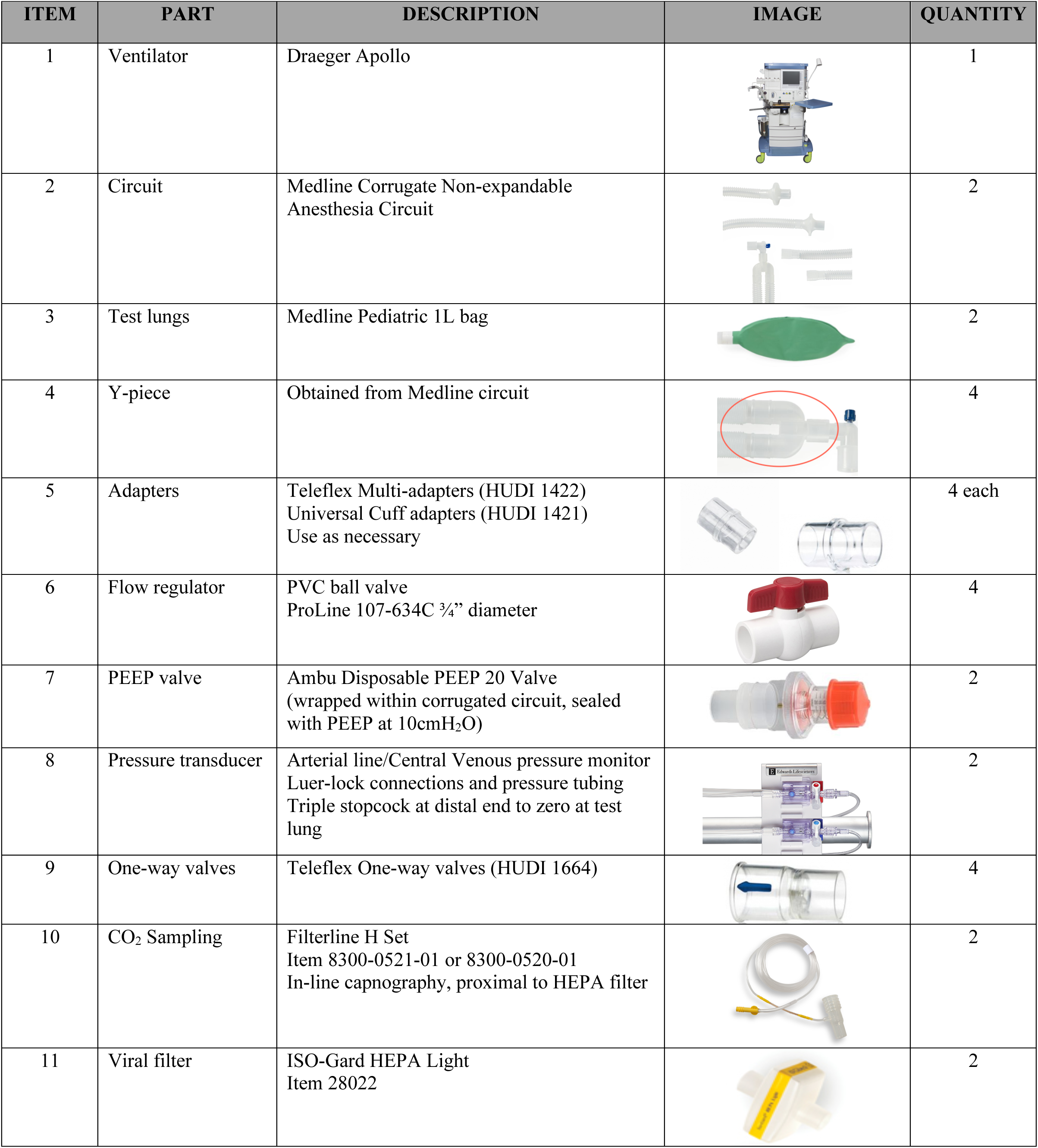
Parts that were utilized in the making of the modified circuit are identified along with model numbers where applicable with images and quantity used. The only product not able to be obtained in the hospital was the ball valve flow regulator. Item numbers correlate with the encircled numbers in the schematic displayed in Figure 1.

Test lungs for Patient A and Patient B were ventilated on pressure control at 12 breaths/minute and an inspiratory/expiratory (I:E) ratio of 1:2. Pressures experienced at each lung for each simulation have been plotted and labeled (Figure 3A and 3B). Of note, the ventilator applies pressure in “cmH_2_O” therefore the pressure transducer’s conventional units (mmHg) have been converted to cmH_2_O (1.36cmH_2_O = 1mmHg).

In Figure 3A, ventilation parameters are identical with no flow restriction. In Figure 3B, PEEP of 10cmH_2_O was set on the ventilator with an additional 10cmH_2_O PEEP placed on Patient A’s expiratory limb via PEEP valve. Patient A’s lung compliance was also significantly decreased by wrapping a rubber tourniquet around the lung. Varying degrees of flow was restricted to both patients by partly closing respective flow regulators. The above conditions were also tested in volume control.

**Figure 3:**
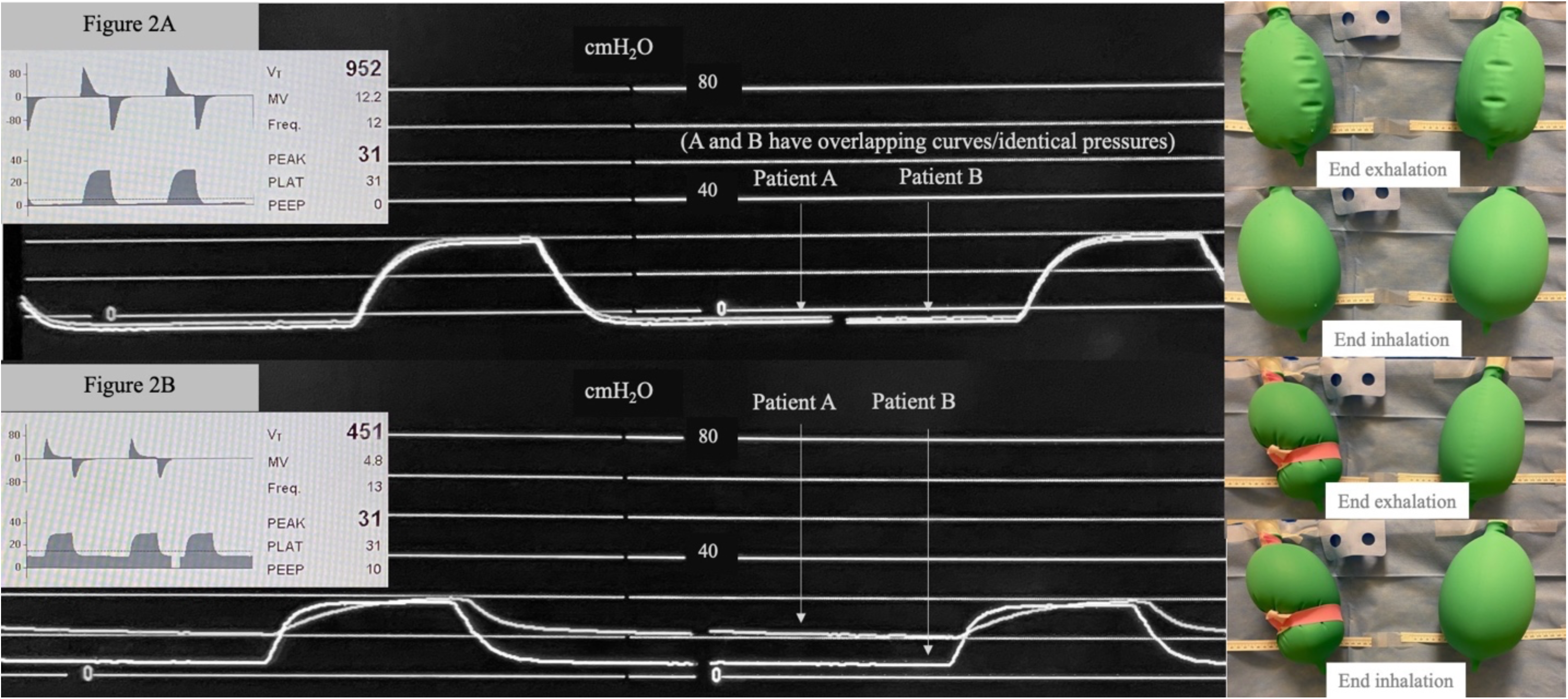
Two test lungs are ventilated under two separate simulations (A) and (B). Ventilator flow and pressure readouts along with combined tidal volume and ventilator PEEP settings are seen in the inset graphs. Patient-labeled pressure curves demonstrate distal pressures experienced by individual lungs throughout the respiratory cycle. The images of test lungs provide qualitative observation of inhalation and exhalation in different simulations. **(A):** Two simulated patients with equal lung compliance are ventilated with identical settings (peak inspiratory pressures, PEEP). With no dialed in flow restriction, both patients appear to receive equal tidal volume. **(B):** Two patients with different lung compliance are ventilated with different settings. Patient A (simulating ARDS) receives protective ventilation (low tidal volume and high PEEP) while patient B (simulating less severe disease), is ventilated with larger tidal volume and lower PEEP.

## RESULTS

When ventilatory parameters were identical, inspiratory pressures generated a combined tidal volume of ~950cc, test lungs qualitatively inflated and deflated identically, and lung pressures were identical graphically (~22mmHg = ~30 cmH_2_O) (Figure 3A; Supplementary video). When flows were restricted to lungs of different compliance in the setting of different PEEP, total delivered tidal volume decreased to ~450cc. Test lungs of Patient A experienced higher end expiratory pressures than those of Patient B as evident by their varying baselines. Their pressure curves were also significantly different from each other. On qualitative observation, Patient A’s test lungs appeared to experience an overall absolute decrease in tidal volume than those of Patient B (Figure 3B; Supplementary video). The peak pressures experienced by lungs of Patient A or B never exceeded the set inspiratory pressure at any point in the pressure control simulations.

## DISCUSSION

Our results demonstrate that ventilation can be manipulated to qualitatively deliver individually tailored tidal volumes in the setting of varying PEEP and compliance requirements in pressure control. As expected, when this was attempted in volume control, flow restriction to one lung led to imbalanced ventilation and over-sized tidal volumes delivered to the other lung. However, when set in pressure control mode, we successfully converted one ventilator into two. Such a force multiplier could immediately save lives, as currently there are ongoing ethical discussions on how to ration this dwindling resource.

Our study is subject to several limitations. First, we performed the experiment using a modern anesthesia machine ventilator rather than an intensive care unit (ICU) ventilator. However, many such machines stand idle or are underutilized, allowing our intervention to be a major force multiplier. Second, lacking a flowmeter (given supply shortfalls), our design set-up could not accurately report split tidal volumes in real time. However, such flowmeters are generally accessible and will likely be available again soon for clinical use. Third, certain ventilation parameters could not be split, namely, Fraction of inspired oxygen (FiO_2_), respiratory rate, and inspiratory time. Future improvements of our proposed design will likely overcome some of these shortfalls. Lastly, some equipment (e.g. ball-valve) was utilized in a manner for which they were not originally designed. However, such parts are commonly used in other industries (e.g. plumbing) and are engineered to withstand fluid pressures far greater than those encountered in human ventilation. In all, these results support the careful administration of multi-ventilation as a viable last-resort alternative in a controlled and supervised environment.

## Legend for Supplementary Video File

**Multi-Ventilation with varying tidal volume and PEEP**. This video demonstrates two multi-ventilation scenarios. In the first scenario, two “patient” lungs of identical compliance are subjected to identical ventilation parameters on pressure control, yielding qualitatively even distribution of total tidal volume. In the second scenario, lungs of different compliance experience different inspiratory pressures and PEEP, yielding independently controlled tidal volumes. Patient A experiences high PEEP and a relatively low tidal volume while Patient B experiences low PEEP and a relatively high tidal volume.

## Data Availability

All data is included in the manuscript

## Glossary of Terms

COVID-19: Novel coronavirus disease of 2019
SARS-CoV-2: Severe acute respiratory syndrome coronavirus
PEEP: Positive end expiratory pressure
IHME: Institute for health metrics and evaluation
ARDS: Acute respiratory distress syndrome
I:E ratio: Inspiratory: expiratory ratio
ICU: Intensive care unit
FiO_2_: Fraction of inspired oxygen

